# Allostatic load in early adolescence: gene / environment contributions and relevance for mental health

**DOI:** 10.1101/2023.10.27.23297674

**Authors:** Kevin W. Hoffman, Kate T. Tran, Tyler M. Moore, Mārtiņš M. Gataviņš, Elina Visoki, Grace E. DiDomenico, Laura M. Schultz, Laura Almasy, Matthew R. Hayes, Nikolaos P. Daskalakis, Ran Barzilay

**Author notes:** Correspondence: Corresponding author: 10 Gates, 3400 Spruce, University of Pennsylvania, Philadelphia, PA 19104.

## Abstract

**Background:** Allostatic load is the cumulative “wear and tear” on the body due to chronic adversity. We aimed to test poly-environmental (exposomic) and polygenic contributions to allostatic load and their combined contribution to early adolescent mental health.

**Methods:** We analyzed data on N = 5,035 diverse youth (mean age 12) from the Adolescent Brain Cognitive Development Study (ABCD). Using dimensionality reduction method, we calculated and overall allostatic load score (AL) using body mass index [BMI], waist circumference, blood pressure, blood glycemia, blood cholesterol, and salivary DHEA. Childhood exposomic risk was quantified using multi-level environmental exposures before age 11. Genetic risk was quantified using polygenic risk scores (PRS) for metabolic system susceptibility (type 2 diabetes [T2D]) and stress-related psychiatric disease (major depressive disorder [MDD]). We used linear mixed effects models to test main, additive, and interactive effects of exposomic and polygenic risk (independent variables) on AL (dependent variable). Mediation models tested the mediating role of AL on the pathway from exposomic and polygenic risk to youth mental health. Models adjusted for demographics and genetic principal components.

**Results:** We observed disparities in AL with non-Hispanic White youth having significantly lower AL compared to Hispanic and Non-Hispanic Black youth. In the diverse sample, childhood exposomic burden was associated with AL in adolescence (beta=0.25, 95%CI 0.22-0.29, P<.001). In European ancestry participants (*n*=2,928), polygenic risk of both T2D and depression was associated with AL (T2D-PRS beta=0.11, 95%CI 0.07-0.14, P<.001; MDD-PRS beta=0.05, 95%CI 0.02-0.09, P=.003). Both polygenic scores showed significant interaction with exposomic risk such that, with greater polygenic risk, the association between exposome and AL was stronger. AL partly mediated the pathway to youth mental health from exposomic risk and from MDD-PRS, and fully mediated the pathway from T2D-PRS.

**Conclusions:** AL can be quantified in youth using anthropometric and biological measures and is mapped to exposomic and polygenic risk. Main and interactive environmental and genetic effects support a diathesis-stress model. Findings suggest that both environmental and genetic risk be considered when modeling stress-related health conditions.

## Introduction

Allostatic load (AL) is the ‘wear-and-tear’ physiological systems in response to managing cumulative chronic stress.^1–3^ AL involves changes in the metabolic, endocrine, cardiovascular, immune, gastrointestinal, and both autonomic and central nervous systems.^4,5^ AL most commonly affects levels of biochemical stress mediators (e.g., cortisol, dehydroepiandrosterone [DHEA]), metabolic markers (e.g., blood glucose and cholesterol levels), and physiologic measures (e.g., blood pressure, body mass index [BMI]).^6^ Most research on AL in humans has focused on adult populations and has shown compelling evidence that ties AL to many negative outcomes involving mental health,^7–12^ physical health,^1,13–18^ and all-cause mortality.^19–22^ Given that differences in AL reflect differences in exposure to stressors, which are tightly associated with social determinants of health,^13,20,23^ it has been suggested that AL may be key to understanding health disparities.^24,25^ Quantification of AL early in the lifespan is critical given the growing recognition that mental and physical disease trajectories and their related health disparities emerge in childhood and adolescence.^26–29^ However, there is no consensus on measurement and operationalization of AL among youth.^8,30–35^

AL emerges as a cost of the chronic activation of stress-biology pathways in response to environmental adversity.^4,36,37^ Upon exposure to stress, the human body responds with higher states of arousal in adaptation to the future demands of the environment, in a state termed *allostasis,* with the ultimate physiological goal to maintain reinstate stability through flexible regulation.^38^ Chronic exposures to stress result in repeated, potentially prolonged, overstimulating, and energetically costly allostasis events, leading to AL.^36,39,40^ In addition to the role of environmental stress exposures in AL, metabolic processes and conditions that relate, such as obesity, have strong genetic underpinning.^41,42^ The combined contribution of environmental stress exposures and genetic susceptibility can be integrated in the conceptual framework of the diathesis-stress model, whereby genetic predisposition and environmental stress exposures interact to determine AL outcome.^43,44^ Evidence in adults has shown that genetic contributions to AL are modest in comparison to the high contribution of environmental factors to AL.^45^ However, the sparse data on genetic (mostly single variant) and environmental contribution to AL in adolescents highlight the possibility for gene-environment interaction effects in the emergence of AL.^46,47^ Notably, gene-environment studies are complicated due to the large genotyped sample sizes needed for adequate power and the methodological challenge of measuring environmental exposures (i.e., exposome). Converging advances in (*i*) statistical genetics that enable the use of generalizable polygenic risk models^48^; (*ii*) statistical methods that quantify aggregate environmental burden,^49^ and (*iii*) the availability of large-scale data of genotyped youth cohorts with detailed phenotypic measures, create opportunities to unravel genetic and exposomic mechanisms in the development of AL.^50^

Here, we leverage multimodal data on thousands of genotyped youths from the Adolescent Brain Cognitive Development^SM^ Study (ABCD Study®) who were followed from late childhood into early adolescence. The ABCD Study emphasized collection of multi-level environmental data that have enabled us to characterize the exposome.^49^ It includes anthropometric and physiological measures, as well as biomarkers that relate to AL as well as mental health measures. We aimed to quantify AL in early adolescence, use exposomic and polygenic risk models to validate AL measurements and test G X E interaction, and determine the mediating role of AL in the pathways from exposomic and polygenic burden to early adolescence mental health. We hypothesized that exposome and polygenic risk scores of metabolic and of stress-related diseases will be additively associated with AL, and that AL will mediate the longitudinal relationship between exposomic and polygenic risk and adolescent mental health burden. We further hypothesized that, because minority youth experience greater adversity,^51^ there will be marked disparities in AL in early adolescence.

## Methods

### Participants

ABCD Study participants (N=11,876) were enrolled at age 9-10 through school systems at 21 sites across the US, with a catchment area encompassing over 20% of the entire US population in this age group.^52^ The current analysis included youth who participated in the third assessment wave of the study (n=10,409, mean age 12). Main analyses were conducted on n=5,035 participants that provided at least one biological sample (blood or saliva) and sensitivity analyses on participants that had at least one measure (anthropometric or biological) that relates to allostatic load (n=8,819). We utilized the ABCD data release 4.0, which includes data collected from the first three assessment waves (age range 9-13). All participants gave assent. Parents/caregivers signed informed consent. The ABCD protocol was approved by the University of California, San Diego Institutional Review Board (IRB) and was exempted from a full review by the University of Pennsylvania IRB.

### Overall study design

We identified measures collected in the ABCD Study that relate to allostatic load (AL) in adolescence^53^ and calculated an overall AL score. We then tested associations of exposomic and polygenic risk scores with the AL score. Additionally, we tested association of AL with psychopathology and tested the mediating role of AL on the pathway from exposomic and polygenic risk burden to psychopathology. **Figure 1** provides a visual representation of the study design.

**Figure 1.**
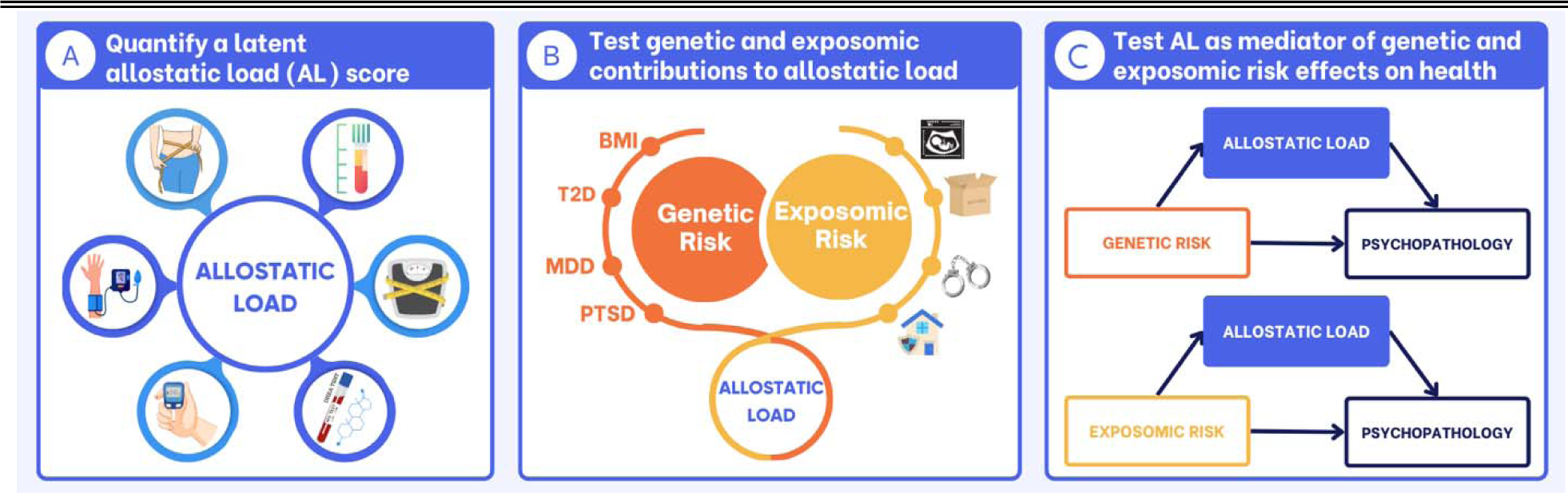
Conceptual study design. **(A)** Using principal component analysis, we quantified a latent allostatic load score (AL). We used BMI, waist circumference, salivary (dehydroepiandrosterone) DHEA levels, blood pressure, and blood levels of hemoglobin A1C and cholesterol to derive the score. **(B)** Then, we used linear mixed effects regression models to test the main and interactive associations of exposomic and polygenic risk scores with AL. Exposomic risk was quantified using a single exposomic factor score that reduces dimensionality of all environmental exposures captured in ABCD by age 11, as we previously described.^49^ Polygenic risk was quantified using polygenic risk scores (PRS) for type 2 diabetes (T2D), and major depressive disorder (MDD). BMI and PTSD PRS were used in sensitivity analyses. **(C)** Lastly, we tested the mediating role of AL on the pathway from exposomic and polygenic risk to youth psychopathology (self-reported measured with the Brief Problem Monitor [BPM] and the parent-reported measured with the Child Behavioral Checklist [CBCL]) scores.

### Quantifying Childhood Exposomic burden

We used a single measure to capture cumulative adverse exposomic burden in childhood (by age 11). This measure was calculated following dimensionality reduction of 348 environmental variables collected by the second ABCD assessment wave, as we previously described.^49^ Briefly, we conducted iterative exploratory factor analyses on environmental measures and identified six domains of environment^54^ (*household adversity* factor, *neighborhood environment* factor, *day-to-day experiences* factor, *state environment* factor, *family values* factor, and *pregnancy/birth complications* factor). Thereafter, we estimated a bifactor model^55^ that generated a general adverse exposome factor and six orthogonal subfactors. The general exposome factor captures shared variance of adverse environment, and we therefore used this measure to quantify overall childhood exposomic burden.

### Quantifying Polygenic risk

We calculated PRS for type-2 diabetes (T2D-PRS) and major depressive disorder (MDD-PRS) using computational pipelines previously described by our group.^56,57^ We used summary statistics from T2D^58,59^ and MDD^60,61^ GWAS. PRS were calculated for European-ancestry (EUR) and African-ancestry (AFR) participants using EUR and AFR GWAS data, respectively. Briefly, PRS-CS (PRS using SNP effect sizes under continuous shrinkage)^62^ was used to infer posterior effect sizes of SNPs in the dataset that overlapped with the GWAS summary statistics, and an external 1,000 Genomes linkage disequilibrium (LD) panel matched to the ancestry group used for the GWAS. Raw PRS were produced by PLINK 1.9 and then standardized in R. The first 10 genetic principal components were regressed out of the standardized PRS. We also used PRS for obesity (BMI-PRS, available only for EUR participants^63^) and post-traumatic stress disorder (PTSD-PRS^64)^ in sensitivity analyses.

### Quantifying Allostatic Load

We modeled AL through a factor analysis of the following eight variables identified in the literature as relevant to allostatic load in adolescence^53^ : BMI, waist circumference, systolic and diastolic blood pressure, blood HDL and non-HDL cholesterol, blood hemoglobin A1C or glycated hemoglobin (HbA1c), and salivary dehydroepiandrosterone (DHEA). Distributions of all eight variables were winsorized at 1%. Structure of allostatic load variables was first determined by exploratory factor analysis (EFA) using least-squares extraction and oblimin rotation. The number of factors to extract was determined by interpretability and the minimum sample-size adjusted Bayesian information criterion (BIC).^65^

To obtain an overall estimate of allostatic load (which we refer to as “AL score” throughout the manuscript), we estimated a bifactor model^66^ through a confirmatory factor analysis (CFA) in Mplus using the robust maximum likelihood estimator. This bifactor model includes variables that load on both a specific factor (e.g., systolic and diastolic pressure on “blood pressure”) and a general (overall) factor. Fit of the model was judged based on the comparative fit index (CFI), root mean-square error of approximation (RMSEA), and standardized root mean-square residual (SRMR). AL scores were estimated for all ABCD participants using all available data. That is, factor score calculation in the presence of missing data in Mplus approximates what the value would be if the data were complete, as opposed to, for example, using mean-imputation on missing data before calculating scores. To minimize measurement error in our main analyses, we only included participants who had at least one biological (blood or saliva) measurement in addition to their physical measurements (N=5,035). Sensitivity analyses included all participants that had at lease one of the eight AL-related measurements (N=8,819).

## Statistical Analysis

The analytic plan and hypotheses regarding association of exposomic, polygenic, and psychopathology measures with AL were registered prior to analysis in May 2023 (https://osf.io/jktg3/?view_only=6b9b36b1c7234e8685719246ebc42bd1). Analyses were conducted between June and October 2023 using R statistical software version 4.1.2 (R Project for Statistical Computing).

### Comparison of AL across sex, race and ethnic subgroups

To examine group differences in AL, we first compared AL scores across subgroups based on sex, race, and ethnicity. Nonparametric tests were used with their corresponding effect size indices, specifically, for the comparison across three race/ethnicity groups, we used Kruskal-Wallis (^2^) and, for pairwise comparison between the race/ethnicity groups, Dunn’s Kruskal-Wallis multiple comparisons with Holm-adjusted p-values. For two-group comparisons (binary sex assigned at birth), we used Mann-Whitney tests (Glass rank biserial coefficient 1^). To examine differential associations of exposomic burden with AL across subgroups, we tested the interaction between exposomic risk scores and sex, race, and ethnicity in association with AL in linear mixed effects models (described below). A significant interaction would suggest that the association between exposomic burden and AL is stronger in a certain subgroup.

### Linear mixed-effects models

To test exposomic and genomic contributions to AL, we used linear mixed-effects models (which we refer to as “mixed models”) with AL score as the dependent variable and childhood exposomic burden or polygenic scores as the independent variables. We estimated 5 mixed models. The basic model tested the association between demographics (age and sex) and AL score (Model 1). We then added childhood exposomic burden to the model (Model 2). We further tested the association between polygenic risk (T2D-PRS or MDD-PRS) and AL score (Model 3). Model 4 included both exposomic and polygenic risk to test their additive association with AL. Lastly, we added the exposomic X polygenic interaction term (Model 5).

To determine the additive contribution of exposomic and polygenic scores to the explanatory models, we compared the variance explained (Nakagawa *R*^2^ ^67^) using the chi-square difference test between nested pairs of models and reported FDR-adjusted p-values.

To test association of AL with psychopathology, we estimated mixed models with either self-report (Brief Problem Monitor [BPM] score ^68^) or parent-report (Child Behavioral Checklist [CBCL] score ^69^) overall psychopathology as the dependent variable and AL as the independent variable.

All mixed models took the hierarchical nature of the ABCD data into account by including a 3-level hierarchy where participants were nested according to family and then were nested according to the research site. All models included random intercepts for site and family.

### Mediation analysis

To assess the mediating contribution of AL to environmental and genetic pathways contributing to adolescent psychopathology, we estimated mediation models in 500 bootstrapped samples (bootstrapped to estimate confidence intervals of standardized effects) using the lavaan R package.^70^ We tested the mediating role of AL to psychopathology in three different pathways: from (i) childhood exposomic burden, (ii) T2D-PRS, and (iii) MDD-PRS. Mediation models included the demographic covariates described above.

## Missing Data

For the measurement model of AL, we utilized pairwise deletion, whereby participants were included even if they had some items missing. With the model estimated using pairwise deletion, scores were calculated using all available data per person. Note that scores calculated in Mplus do not use imputation, but rather, approximate using available data what the score would be with complete data. This means scores calculated in the presence of missing data are not “weighted toward the mean,” as would be the case if mean- or median-imputation were used on missing values before calculating scores.

For all mixed models, we utilized listwise deletion of missing data. The proportions of missing data for all measures used in the study sample are provided in **Supplementary Table 1**.

## Sensitivity Analyses

To assess potential unexpected effects of missing data, we included two additional analyses where we tested association of exposomic burden with AL scores in:

1. Only ABCD Study participants who did not have any missing data on the measures we included in the AL measurement model (n=284).
2. All ABCD Study participants who participated in the third assessment wave and had at least one measure that relates to AL (N=8,819), not only those with biological data as in main analyses.

To assess specificity of our findings regarding genomic contributions to AL, we substituted T2D-PRS with BMI-PRS and MDD-PRS with PTSD-PRS in all models testing genomic associations with AL.

## Results

### Allostatic load in ABCD

An EFA of the eight measures that relate to allostatic load revealed four factors including Weight (BMI and waist circumference), Blood pressure (systolic and diastolic), Cholesterol (non-HDL and HDL cholesterol) and Other, which included the remaining two biomarkers (HBA1C and DHEA). Note that DHEA showed significant loadings on both the Weight (0.6) and Other (-0.46) factors, but we chose to include it in Other based on theory, as DHEA is not a weight measurement and because both salivary DHEA and HBA1c levels can be conceptualized as stress-related biomarkers.^71,72^ The detailed EFA loadings of the variables are shown in **Supplementary Table 2**.

To estimate a general AL score, we fit a bifactor model.^55^ This model included an overall AL factor in addition to four orthogonal subfactors as revealed in the EFA described above. **Figure 2A** shows the results of the bifactor analysis with the loadings of all items. Fit of the model was acceptable,^73^ with a CFI of 0.997, RMSEA of 0.017 and SRMR of 0.029. BMI and waist circumference were the strongest loading items to the general AL score (0.81 and 0.75, respectively). AL was modestly correlated with age (r=.12, P<.001). **Figure 2B** includes all correlations among AL related items in study participants.

**Figure 2.**
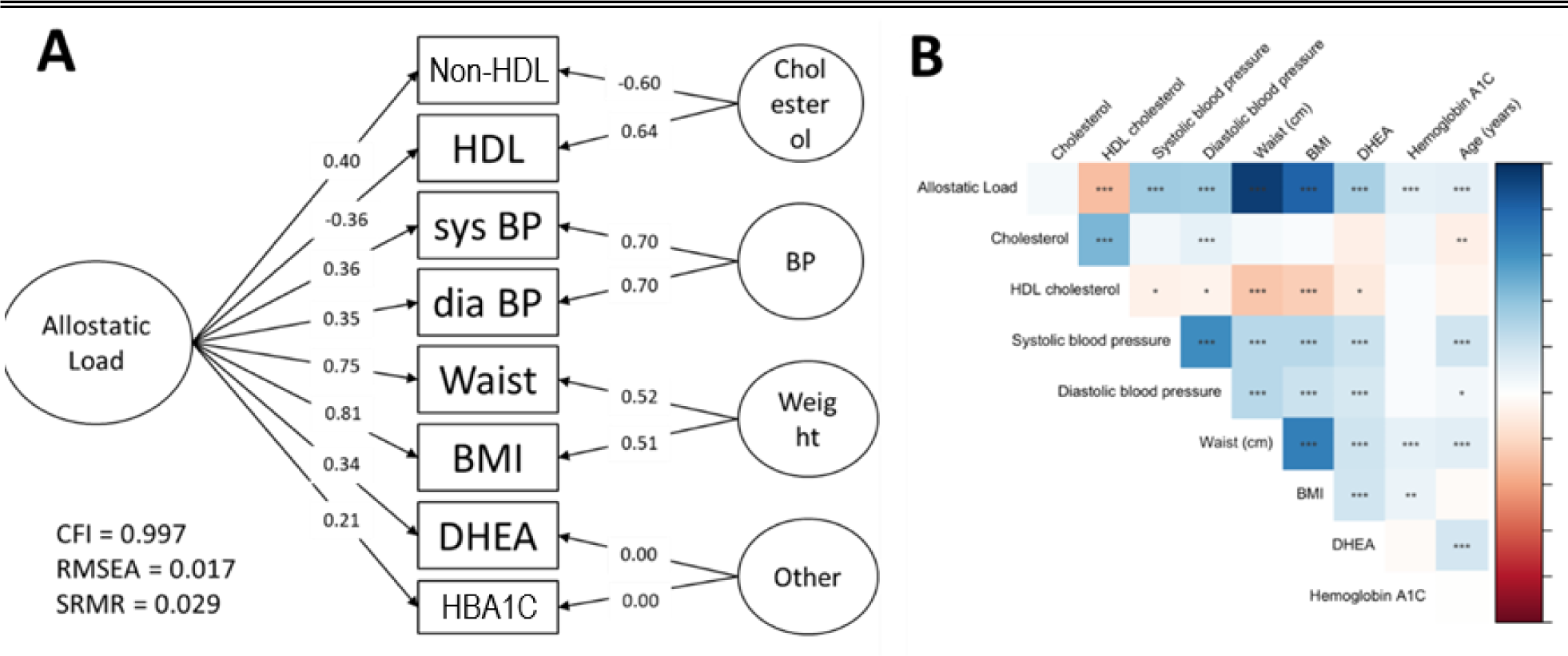
Allostatic load measurement in ABCD Study. **(A)** The EFA performed before the bifactor analysis resulted in 4 factors, which determined the 4-factor configuration of the bifactor model (right-hand side of panel A), with loadings to each factor shown on the arrows pointing to individual measures. Main allostatic load factor loadings are shown on each arrow from allostatic load to individual measure. Measures of model fit (CFI, RMSEA, SRMR) are provided on the bottom left. **(B)** Correlation heatmap of the individual variables, age, and the general allostatic load score. *P<.05; **P<.01; ***P<.001. Abbreviations: HDL, high density lipoprotein; sys BP, systolic blood pressure; dia BP, diastolic blood pressure; BMI, body mass index; DHEA, dehydroepiandrosterone; HBA1c, glycated hemoglobin; CFI, comparative fit index; RMSEA, root mean-square error of approximation; SRMR, standardized root mean-square residual.

### Association of childhood exposomic burden with overall AL

Based on the conceptualization that AL is driven by cumulative stress exposure, we tested the longitudinal association of overall childhood exposomic burden, modeled using a single overall adverse exposome calculated at age 11^49^ with AL in the diverse study sample (N=5,035, see **Table 1** for detailed demographic characteristics). Exposomic burden was significantly associated with AL a year later (beta=0.25, 95%CI 0.22-0.29, P<.001; **Figure 3A**). This association was attenuated but remained significant after adjusting for age, sex, race, ethnicity, household income, and parental education (beta=0.14, 95%CI 0.10-0.19, P<.001; Supplemental Table 3).

**Table 1.** Characteristics of ABCD Study youth with anthropometric and biological measures included in this study.

|  | STUDY POPULATION (N=5,035<sup>^</sup>) |
| --- | --- |
| Age, years (mean (SD)) | 11.97 (0.64) |
| Female sex assigned at birth, n (%) | 2,404 (47.75) |
| White, n (%) | 4,026 (79.96) |
| Black, n (%) | 814 (16.17) |
| American Indian or Alaska Native, n (%) | 186 (3.69) |
| Native Hawaiian or Other Pacific Islander, n (%) | 41 (0.81) |
| Asian, n (%) | 292 (5.80) |
| Other, n (%) | 268 (5.32) |
| Mixed race, n (%) | 555 (11.02) |
| Hispanic/Latinx, n (%) | 944 (18.75) |

<sup>^</sup>Note that sample size of main analysis was limited to participants who provided at least one biological sample that related to allostatic load (blood HBA1C, blood cholesterol or salivary DHEA).

**Figure 3.**
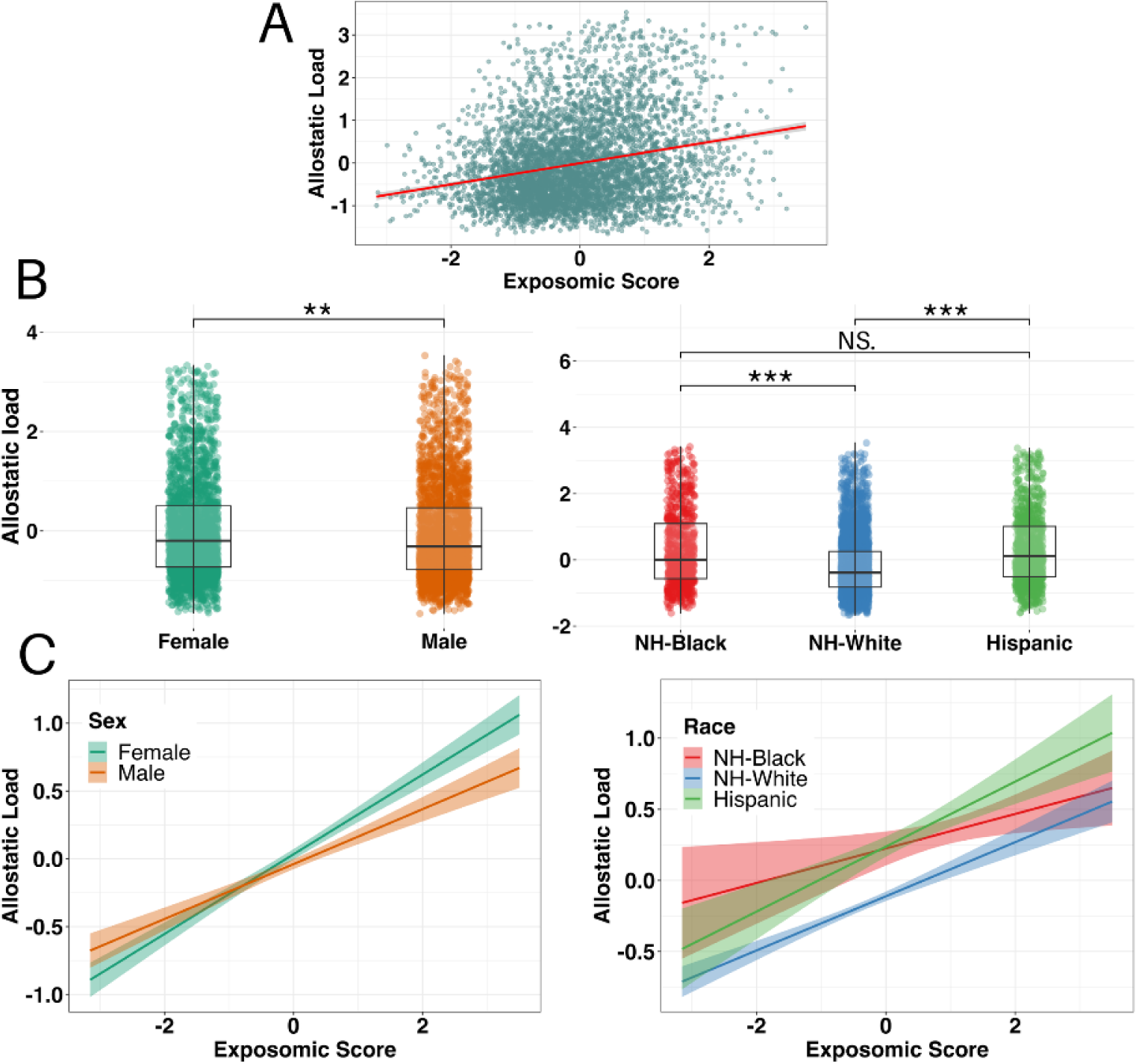
Association of exposomic burden with allostatic load and demographic differences. **(A)** Scatter plot depicting the association of exposomic score at the second ABCD Study assessment wave (age 11) as independent variable and allostatic load at the third assessment wave (age 12) as dependent variable. **(B)** Comparison of AL scores by demographic groups – (left) comparison by sex assigned at birth, (right) comparison by racial and ethnic grouping. Each dot represents a participant, boxplots show median, with the box denotes interquartile ranges (bottom indicated Q1, top – Q3) and the whiskers extend to the minimum and maximum values. Asterisks denote significance (** corresponds to p-value < 0.01 and *** to p-value < 0.001). **(C)** Regression lines depicting moderation of demographic characteristics – (left) sex assigned at birth, (right) racial and ethnic grouping-on the association of exposomic score at the second assessment wave (age 11) as independent variable and allostatic load at the third assessment wave (age 12) as dependent variable. Shaded bands denote 95% pointwise confidence intervals. Abbreviations: NH-Black, non-Hispanic Black youth; NH-White, non-Hispanic White youth.

### Differences in allostatic load across subgroups

There were small but significant sex differences (female greater than male participants, median score -0.20 vs. -0.31, respectively, P=.002), and significant racial and ethnic disparities in AL (**Figure 3B**). Non-Hispanic White youth had significantly lower AL (median=-0.39) compared to Hispanic (median=0.12, P<.001) and non-Hispanic (NH) Black youth (median=0.001, P<.001). We did not observe difference in AL between Hispanic and NH-Black youth (P=.50).

### Differential associations of exposomic burden with allostatic load across subgroups

The association between exposomic burden and AL was stronger among females compared to males (significant exposome by sex interaction in association with AL, coefficient = -0.09; 95% CI = -0.15 – -0.04, P=.001; **Figure 3C**). There were no differential associations of exposomic burden with AL across race or ethnicity (exposome by Black race interaction P=.07; exposome by Hispanic ethnicity interaction P=.60; **Figure 3C**).

### Polygenic contributions to allostatic load

We tested associations of PRSs representing two domains that we hypothesized are relevant for AL: genetic metabolic susceptibility, modeled with T2D-PRS, and genetic stress susceptibility, modeled with MDD-PRS. In EUR participants (n=2,928), both PRSs were significantly associated with AL (T2D-PRS beta=0.11, 95%CI 0.07-0.14, P<.001; MDD-PRS beta=0.05, 95%CI 0.02-0.09, P=.003; **Supplementary Table 4**). We did not observe significant associations of these PRSs with AL in the AFR participants (n=630) (**Supplementary Table 5**).

### Combined polygenic and exposomic contributions to allostatic load

We tested additive and interactive associations of exposomic and polygenic burden with AL in EUR ancestry participants (**Supplementary Table 4)**. Main effects remained unchanged for both exposomic and polygenic scores for T2D and MDD. While the model with exposomic burden explained 6.0% of the variance in AL, addition of T2D-PRS significantly increased variance explained in AL to 6.97% (Chi-squared P<.001). Addition of MDD-PRS to the model with exposomic burden did not have significant additive effect over variance explained (6.12% vs. 6.0%, respectively, Chi-squared P=.053).

Both PRSs showed significant G X E interactions in association with AL, such that with greater polygenic risk of either T2D-PRS or MDD-PRS, the association between exposomic burden and AL was stronger (T2D-PRS X exposomic burden interaction P=.021; MDD-PRS X exposomic burden interaction P=.045; **Figure 4**, **Supplemental Table 4).**

**Figure 4.**
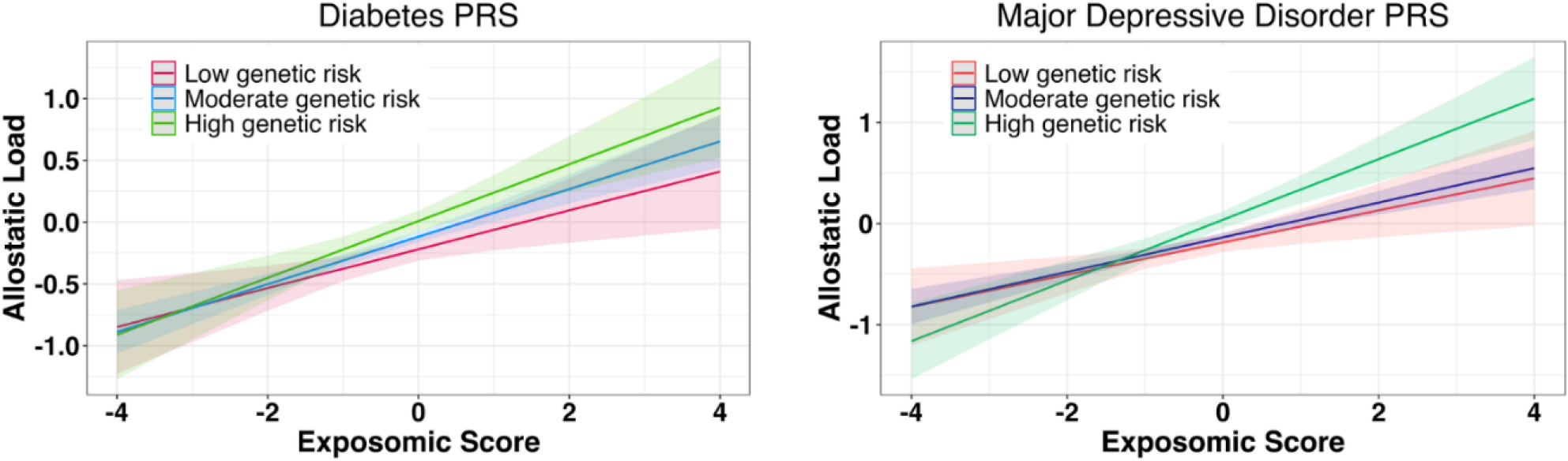
Gene-environment interaction in Allostatic Load. Regression lines depicting moderation of T2D-PRS (left) and MDD-PRS (right) on the association of exposomic score at the second ABCD Study assessment wave (age 11) as independent variable and allostatic load at the third assessment wave (age 12) as dependent variable in European ancestry participants. Both interaction terms were statistically significant (T2D-PRS exposomic burden interaction P=.021; MDD-PRS exposomic burden interaction P=.045). Shaded bands denote 95% pointwise confidence intervals. Abbreviations: T2D, Type 2 diabetes; MDD, major depressive disorder; PRS, polygenic risk score.

### Association of allostatic load with psychopathology

We next examined the cross-sectional associations between AL and psychopathology. AL was associated with both self- and parent-reported symptom burden with a small but statistically significant effect size, adjusting for age, sex, race, ethnicity, household income and parental education (for self-report standardized BPM total scores, effect =0.04, 95%CI=0.01-0.08, P=.008; for parent report standardized CBCL total score, effect=0.08, 95%CI=0.05-0.10, P<.001; **Supplementary Table 6**).

### Mediation effects of AL on the pathway from exposomic and polygenic risk to psychopathology

We finally tested the hypothesis that AL mediates the pathway from exposomic and polygenic risk to psychopathology. For the exposomic pathway, we found that AL partly mediated the path from exposomic risk to parent-reported psychopathology (P = .006 for indirect pathway through AL), with no significant mediation effect on the path to self-reported psychopathology (P=.20; **Figure 5A** and **Supplemental Table 7)**.

**Figure 5.**
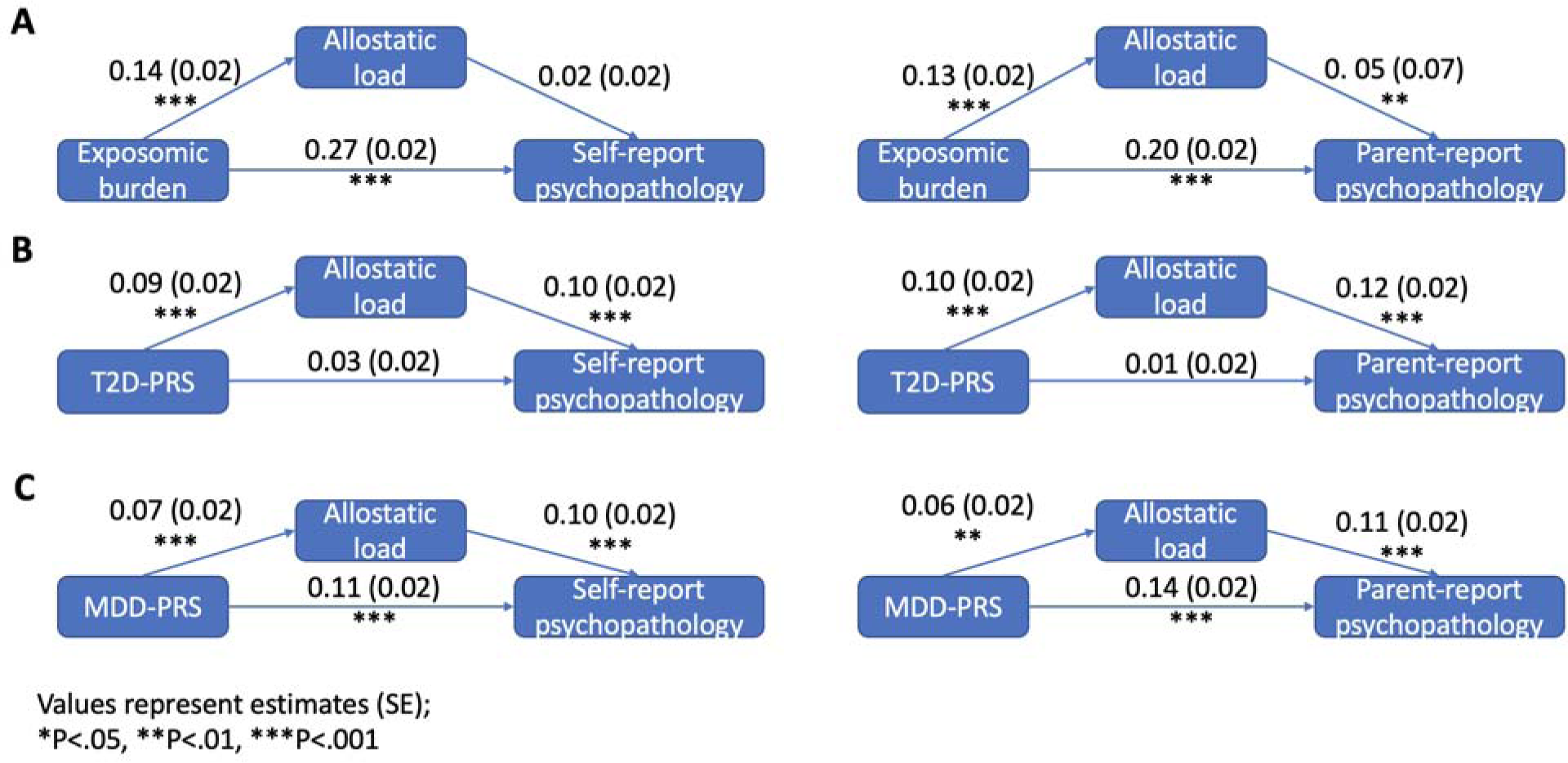
Mediation models. We tested the mediating effect of allostatic load between **(A)** exposomic burden, **(B)** T2D PRS, and **(C)** MDD-PRS and psychopathology. Panel **(a)** (exposomic pathway) includes a diverse population (n=4,043 and n=4,540 for self and parent report, respectively), and panel **(B)** and **(C** (polygenic pathways) include European ancestry participants (n=2,652 and n=2,928 for self and parent report, respectively). All variables are z-scored. Values on the arrows represent estimated beta coefficient values with standard error values in parentheses.

For metabolic polygenic risk in EUR participants, AL fully mediated the path from T2D-PRS to parent-reported psychopathology (P<.001 for the indirect pathway through AL, P=.22 for the total path from T2D-PRS to parent-reported psychopathology, **Figure 5B** and **Supplemental Table 8).** AL partly mediated the path from T2D-PRS to self-reported psychopathology (P<.001 for the indirect pathway through AL, P=.04 for the total path from T2D-PRS to self-reported psychopathology, **Figure 5B** and **Supplemental Table 8).**

For psychiatric polygenic risk in EUR participants, the direct path from MDD-PRS to self- and parent-reported psychopathology was significant (both P’s<.001), with AL partly mediating the path from MDD-PRS to both parent- and self-reported psychopathology (P=.006 and P=.007 for indirect pathway through AL, respectively; **Figure 5C** and **Supplemental Table 8)**.

We did not observe significant mediation effects for AL on the pathway from polygenic risk to psychopathology in the AFR participants (all indirect pathway P’s>.4; **Supplemental Table 9)**.

### Sensitivity analyses

To assess the potential effect of missing data on our findings, we reran our model testing exposomic contribution to AL in participants with no missing data (n=284) or using all participants who were included in the ABCD study third assessment wave and had at least one measurement related to AL (BMI, waist circumference, blood pressure or any biospecimen; n=8,819). Results remained unchanged in direction and statistical significance for all models testing the univariate association between exposomic score and AL (**Supplementary Tables 10-11**). For the multivariate models adjusting for demographics, results remained similar with the single exception that the association between exposomic burden and AL was no longer significant in the multivariable model (P=.23; **Supplementary Tables 10**).

To assess generalizability of our findings regarding polygenic contributions to AL, we reran our main models substituting T2D-PRS with BMI-PRS (as a PRS representing metabolic sensitivity) or substituting MDD-PRS with PTSD-PRS (as a PRS representing psychiatric stress sensitivity). Results remained largely unchanged in direction and statistical significance (**Supplementary Tables 12-13**). Correlations among PRS is shown in **Supplemental Figure 1**.

## Discussion

In this work, we quantify allostatic load in a large sample of American early adolescent youth and test its relationship with exposomic burden, polygenic risk scores, and mental health. Consistent with previous work in both developing and adult cohorts, we show that AL is associated with environmental stress and mental health outcomes^8,11,74^ and that there are racial and ethnic disparities in AL.^12,20,23,75–79^ In line with our hypotheses, we show that polygenic risk scores of metabolic and psychiatric disorders have additive and interactive effects on the relationship between environmental (exposomic) adversity and AL, supporting a diathesis-stress model.^80^ Using longitudinal mediation modeling, we show that AL functions as a mediator between exposomic and polygenic risk and mental health outcomes even in early adolescence. These findings expand existing literature suggesting a mediating role of AL from childhood adversity to adult mental health^81,82^ and support the hypothesis that AL could be a mechanism that explains health disparities.^24^ Critically, we show evidence of this mechanism at an early age, well before routine monitoring for and expected onset of many chronic medical illnesses.

Using environmental and genetic data, our study validates AL in a large community sample of youth and demonstrates that higher exposomic burden predicts increased allostatic load in the future, which is associated with higher mental health burden. Environmental exposures contribute substantially to all health burden,^83,84^ and allostatic load has been proposed as the mechanistic link of environmental contributions to health outcomes.^24^ In addition to exposomic burden, we show racial and ethnic disparities in AL. Studies have highlighted that AL may be a key biological mediator of the minority stress effects on racial disparities in health and mortality.^20,23,24,76,85,86^ Notably, high allostatic load is associated with both mental and physical health outcomes^1^ and with greater all-cause mortality,^19^ with additive contribution of AL to exposomic burden in explaining mortality.^23,76^ Our results in a young developing population therefore concur with the existing literature in older populations regarding the links between environment and AL and between AL and health outcomes. We suggest that AL measurement may be used as a biomarker of hyperactivation of stress systems and might have translational relevance as an indicator of future disease risk. We suggest that our work on AL in community youth propel research in clinical populations, where AL measurements, alongside other patients’ data (e.g., clinical, genetic, social determinants of health) can help parse heterogeneity of health outcomes and disparities in adolescence.

A key findings of our study is the illumination of gene-environment effects on AL. Studies on G X E interaction are challenging and have resulted in inconsistent results when testing interactions between single genes and single exposures,^46,80,87–90^ highlighting the need for development and application of novel tools that quantify genetic and environmental risk.^91^ Advances of statistical genetics methods now allow calculation of polygenic scores that aggregate genetic susceptibility in a single measure.^92^ Adoption of the exposome framework has allowed us to quantify exposomic burden in a single measure.^49^ Here, we combine these tools and show not only additive polygenic and exposomic effects on AL in early adolescence, but also significant G X E interaction. This finding adds to the literature suggesting that genetic effects on youth health outcomes vary by environment.^80^ The interaction we observed concurs with the diathesis-stress model of disease,^80,93,94^ whereby youth with high polygenic risk showed stronger association between environmental burden and AL. Notably, this observation was consistent when using two different types of polygenic scores with two examples in each: metabolic genetic risk (T2D and BMI) and stress-susceptibility genetic risk, which was estimated using polygenic risk scores for stress-related psychiatric disorders (MDD and PTSD). By using both PRS for metabolic and psychiatric conditions, we highlight the integrative nature of AL as a link between physical and mental health, and as a marker of exposomic burden on complex biological systems.

We found that AL partly mediated the longitudinal association between exposomic burden and youth psychopathology at age 12 (parent-reported). This finding aligns with previous works showing the mediating role of AL on the path from specific exposures, childhood physical and sexual abuse^95,96^ and neighborhood poverty,^97,98^ to mental health burden in older adolescents and adults. Our results also indicate that AL mediates the relationship between genetic susceptibility and mental health burden. First, we show that AL partially mediates the path from polygenic risk of MDD, as a model of stress-related psychiatric condition, and early adolescent mental health burden. This finding suggests that at least some of the genetic risk captured in polygenic scores of depression is mediated by genetic stress-susceptibility captured in AL measurement. Second, we found that AL fully mediates the pathway from metabolic genetic risk, estimated by T2D-PRS, to parent-reported youth psychopathology. This finding expands our understanding of the mechanisms linking physical and mental health susceptibility, as observed by the highly prevalent clinical comorbidities between metabolic conditions and mental health problems.^99,100^ This overlap is already observed in adolescence, manifested for example by the high rates of depression in pediatric obesity (34% more likely to be depressed^101^). Notably, our results on the link between metabolic susceptibility, AL, and mental health burden also relate to the allostatic load energy model.^36^ This model suggests that hypermetabolism, the overuse of energy resources for allostasis at the cost of concurrent essential biological processes, drives accelerated aging through modifications of physiological processes, such as mitochondrial function, especially in early life,^102–104^ which may result in accelerated development at the cost of protracted development.^105–107^ Our findings regarding the full mediation of AL on the path from polygenic risk of T2D to mental health burden, as opposed to the partial mediation of polygenic risk of MDD, may suggest that the effects of trait-like metabolic susceptibility on stress biology outweighs stress-related disease susceptibility effects. As allostatic load is implicated in accelerating developmental processes (e.g., precocious puberty^34^ and accelerated cellular aging processes^108,109^), our results present a key justification for the study and clinical utility of metabolic (and other physiological) system susceptibility in mental health and the necessity for an integration of systems approaches to understanding mental and physical health.^83,110,111^ Together, our mediation analyses point toward allostatic load as a contributor on the causal pathway from converging risks of psychiatric and metabolic predispositions to mental health burden in early adolescence and may suggest AL as a potential target for interventions aimed at improving youth mental health outcomes.^36,112^

Our findings should be interpreted in the context of certain limitations. First, we used eight measures to model AL but some measures important for AL such as stress mediators of the HPA axis (e.g., cortisol) and inflammatory markers^53^ were not available. Second, our sample had significant missingness in allostatic load measurements, which may have biased our AL factor scores to overrepresent the variables with lowest missingness, namely waist and BMI. However, our sensitivity analyses showed consistent findings when using participants with all AL measures and when using participants with more missing data, bolstering confidence in our findings. Third, in our choice of polygenic risk scores, stress susceptibility was represented by MDD-PRS and PTSD-PRS, which have been shown to have limited explanations for their heritability.^113^ This may mean that the PRS for stress susceptibility might represent not the actual gene-linked susceptibility but the genetic correlations.^114^ Further work, integrating biological multi-omics must be used to establish the link between polygenic risk for stress susceptibility and allostatic load.^115,116^ Fourth, while we included genetic analyses in African ancestry participants, their sample sizes were substantially smaller, limiting the statistical power to capture the effects observed in European ancestry youth. An additional challenge is that African ancestry PRS use GWAS data with smaller sample sizes, limiting their generalizability to our present study population. We echo the call to include more diverse populations in future medical genetics research to create reliable and generalizable polygenic risk scores in diverse ancestries.^117^ This work is especially critical in the context of our findings of differential exposures to AL in Black and Hispanic youth. Fifth, we used data from a single study. While the ABCD Study includes data on a large diverse sample ascertained in 21 different sites across the US, external validation should be done in other cohorts (including from outside the US) and in different sociocultural environments to ensure generalizability and reliability of our findings. Moreover, for these findings to have clinical impact, we call for external validation in clinical pediatric populations. Lastly, while we used a longitudinal design to test associations of exposomic and polygenic contributions to AL, we cannot infer causality. We hope that our work will propel more research using causal inference statistical methods^118^ to test causal effects of environmental and genetic factors to AL and their combined causal contribution to later health outcomes. This work will be critical for elucidating disease mechanisms and identifying modifiable treatment targets.

In conclusion, we present evidence for the measurement and validity of allostatic load in early adolescence. AL maps to environmental and genetic burden and shows gene-environment interaction in adolescence in line with the stress diathesis model. Furthermore, we report racial and ethnic disparities in AL evident already at a young age and provide evidence suggesting a mediating role for AL bridging exposomic burden, genetic susceptibility, and mental health outcomes. Future work is needed to address open questions that our findings highlight, such as testing AL in youth pediatric clinical populations with high physical health burden and adding more measurements of AL from other organ systems. We suggest that research integrating allostatic load with measures of environmental stress exposures is critical to inform precision medicine, including precision environmental health frameworks, detection of preclinical disease and treatment of early symptoms, and mitigation of disease complications or improvement of outcomes.^83^

## Data Availability

This study involves secondary analysis of the Adolescent Brain Cognitive Development (ABCD)Study. Access to ABCD Study data can be granted to bone fide researchers upon request from the NIMH data archive (NDA).

https://nda.nih.gov/abcd/

